# Multi-gesture drag-and-drop decoding in a 2D iBCI control task

**DOI:** 10.1101/2024.09.17.24313659

**Authors:** Jacob T. Gusman, Tommy Hosman, Rekha Crawford, Tyler Singer-Clark, Anastasia Kapitonava, Jessica N. Kelemen, Nick Hahn, Jaimie M. Henderson, Leigh R. Hochberg, John D. Simeral, Carlos E. Vargas-Irwin

## Abstract

**Objective:** Intracortical brain-computer interfaces (iBCIs) have demonstrated the ability to enable point and click as well as reach and grasp control for people with tetraplegia. However, few studies have investigated iBCIs during long-duration discrete movements that would enable common computer interactions such as “click-and-hold” or “drag-and-drop”.

**Approach:** Here, we examined the performance of multi-class and binary (attempt/no-attempt) classification of neural activity in the left precentral gyrus of two BrainGate2 clinical trial participants performing hand gestures for 1, 2, and 4 seconds in duration. We then designed a novel “latch decoder” that utilizes parallel multi-class and binary decoding processes and evaluated its performance on data from isolated sustained gesture attempts and a multi-gesture drag-and-drop task.

**Main Results:** Neural activity during sustained gestures revealed a marked decrease in the discriminability of hand gestures sustained beyond 1 second. Compared to standard direct decoding methods, the Latch decoder demonstrated substantial improvement in decoding accuracy for gestures performed independently or in conjunction with simultaneous 2D cursor control

**Significance:** This work highlights the unique neurophysiologic response patterns of sustained gesture attempts in human motor cortex and demonstrates a promising decoding approach that could enable individuals with tetraplegia to intuitively control a wider range of consumer electronics using an iBCI.

## 1. Introduction

Intracortical brain-computer interfaces (iBCIs) utilize neural signals from motor areas of the brain to allow people with tetraplegia to control external devices like computers and robots using imagined or attempted movements of the arm and hand [1–3]. Recent work has shown that individual finger movements [4,5], complex hand gestures [6,7], and even handwriting [8] can be reliably decoded using iBCIs. However, each of these control signals represent transient motor commands that evolve over the course of 1 or 2 seconds and are decoded as discrete events. Many upper extremity actions important to everyday activities require maintaining a specific posture over relatively longer periods of time (holding objects, pressing a button for a desired duration, clicking and dragging with a computer mouse, etc.). How do cortical signals evolve during this type of sustained, static motor command?

Neurons in motor and premotor cortices of nonhuman primates (NHPs) have frequently been characterized as “phasic-tonic”; neurons in these areas tend to fire more robustly at the beginning of an isometric grasp attempt, but less so as the grasp force is maintained over time [9, 10]. While neural activity during initial phasic responses tends to correlate with EMG, sustained tonic activity is only weakly correlated with motor output [10–12]. Furthermore, the tuning of NHP motor cortical neurons to different motor commands can decrease over the course of a sustained isometric hold [13, 14], which would make it more difficult for a neural decoder to reliably differentiate between different sustained actions held over time. Additionally, grasp decoding of sustained holds can be attenuated during concurrent arm translation [15].

To date, the majority of iBCI neural decoding systems circumvent these challenges by employing velocity-based control logic. Attempted movements performed by the participant are mapped to discrete or continuous changes in the state or position of cursors and robotic effectors [2, 16–20]. Recent work has shown that intuitive drag-and-drop control of a cursor could be approximated by detecting transient signals corresponding to the initiation and release of a single hand grasp [21]. Here, we utilize a neural “latch” decoder to accommodate sustained gesture attempts performed in a multi-gesture context. Through this approach, we demonstrate the possibility of offering responsive and consistent decoding of multiple gestures over long hold periods.

## 2. Methods

### 2.1 Participants

Study participants T11 and T5 provided informed consent and were enrolled in the pilot clinical trial of the BrainGate Neural Interface System. Enrollment criteria and details about the trial can be found at http://www.clinicaltrials.gov/ct2/show/NCT00912041. T11 is a man in his 30s with tetraplegia due to a C4 AIS-B spinal cord injury that occurred 11 years prior to enrollment in the trial. T5 was a man in his 70s with tetraplegia due to a C4 AIS-C spinal cord injury that occurred 9 years prior to enrollment in the trial. At the time of this study, T11 had been enrolled in the trial for approximately 1 year, and T5 had been enrolled in the trial for approximately 6.5 years.

Permissions for this study were granted by the US Food and Drug Administration (FDA, Investigational Device Exemption #G090003) and the Institutional Review Boards (IRBs) of Massachusetts General Hospital (#2009P000505, initially approved May 15, 2009, includes ceded review for Stanford University and Brown University) and Providence VA Medical Center (#2011-009, initially approved March 9, 2011). All research sessions were performed at the participants’ place of residence.

### 2.2 Intracortical Recordings

Data were recorded via two 96-channel microelectrode arrays (Blackrock Neurotech) placed chronically in the hand-knob area of the left precentral gyrus (PCG) of each participant. Neural signals from T11 were transmitted wirelessly using two ‘Brown Wireless’ devices (Blackrock Neurotech), whereas neural signals from T5 were acquired using two NeuroPort Patient Cables (Blackrock Neurotech). Previous work showed a negligible difference in signal quality between these two signal transmission strategies [22]. From these recordings, we extracted neural features that included threshold crossings (TX) [23], spike band power (SP) from the 250-5000Hz band (8th order IIR Butterworth), and short-time Fourier transform extracted local field potential (LFP) power across five bands, 0-11 Hz, 12-19 Hz, 20-38 Hz, 39-128 Hz, and 129-250 Hz.

## 3. Latch Decoder Development

### 3.1 Multi-Duration Gesture Study: Gesture Hero Task

To better understand how sustained gestures were represented in the neural recordings from our participants, we asked T11 and T5 to attempt hand gestures for differing durations using the Gesture Hero Task [7]. Like its video game namesake (Guitar Hero, Activision) upcoming movements were instructed by falling boxes with a picture of the cued gesture (Fig. 1A, Video S1*‡*). The participants were asked to attempt and maintain the gesture when the falling box came in contact with the “attempt line” positioned at the lower third of the screen and to relax after the box fell past the attempt line. Therefore, the attempt period was indicated by the amount of time the box intersected with the line. Since boxes fell at a constant speed, the size of the box was directly proportional to the attempt time, which was either 1, 2, or 4 seconds long. The instruction period, the time it took the box to move from the top of the screen to the attempt line, was four seconds. The intertrial period, the time between the box exiting the attempt line and the appearance of the next gesture box, was 1.3 seconds.

**Figure 1.**
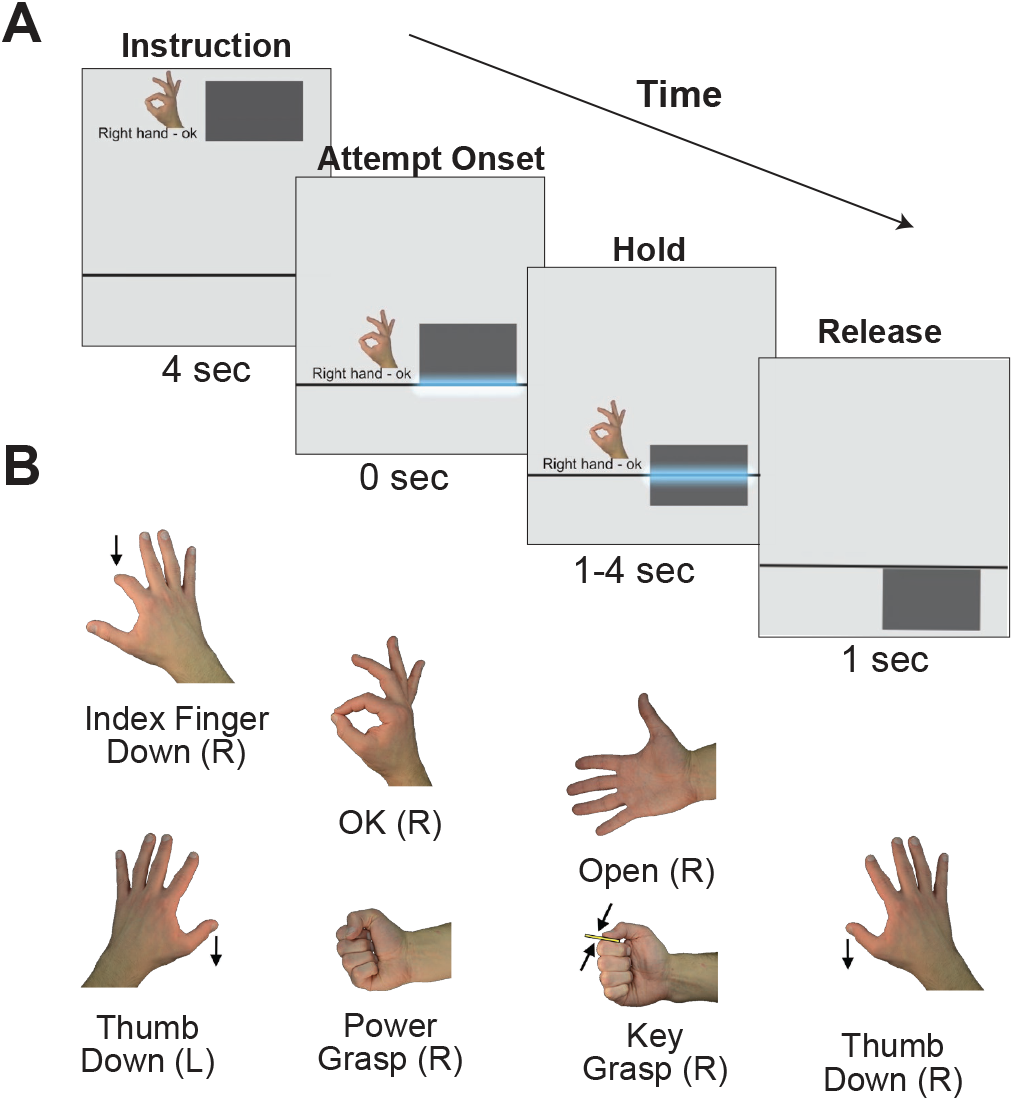
Gesture Hero Task. (A) Gesture Hero Task. Instruction: Boxes with the upcoming gesture fell from the top of the screen to the attempt line (gray line) in four seconds. Attempt Onset: The participant was requested to attempt and hold the gesture when the box contacted the attempt line. Hold: The hold period lasted for 1, 2, or 4 seconds. Release: After the box passed the attempt line, there was a 1.3 second intertrial interval before the next gesture box appeared. (B) Gestures presented in the task. Right hand: Index finger down, OK, Open, Key grasp, Power grasp, Thumb down. Left hand: Thumb down.

T11 was asked to attempt six right-handed gestures (index finger down, power grasp, OK, open, key grasp, thumb down) and one left-handed gesture (thumb down; Fig. 1B). He performed the Gesture Hero Task on two session days, each session consisting of ten data blocks and twenty trials per gesture-duration condition. Due to time constraints, data collection with T5 was limited to using a subset of three gestures - index finger down, power grasp, and OK - in one session with twenty trials per gesture-duration condition.

### 3.2 Gesture Information During Sustained Attempts

Using the dataset from the Gesture Hero task we evaluated the representation of gesture information over the course of the different hold durations. Similar to what was observed among neurons in NHPs [13, 14], gesture-selective activity appears to decrease during isometric holds of greater than 1 second (Fig. 2). For example, whereas about 25% of the neural features (TX and SP) recorded from T11 were gesture selective (i.e. displayed significantly different values across gesture conditions; Kruskal-Wallis test, *p <* 0.001) after 1s holds, 16% of features were gesture selective after 2s holds and 10% of features were gesture selective after 4 second holds (Fig. 2A). Furthermore, by assessing the performance of an LDA classifier over the trial durations it becomes clear that standard LDA-based decoding approaches would not reliably maintain gesture decoding throughout extended attempt periods (Fig. 2B). For example, during a 4 second sustained gesture attempt, a simple LDA classifier would correctly decode about 60% of 300ms data samples centered at the 1s mark after attempt onset, compared to 30% correct decodes at the 4s mark, representing a 50% decrease in classifier performance. However, we found that if we applied LDA classification to the far simpler task of determining whether or not any gesture attempt was being performed (“Attempt Classification”, Fig. 2C), we observed a less drastic drop off in decoding accuracy (11.5% decrease from 1s to 4s).

**Figure 2.**
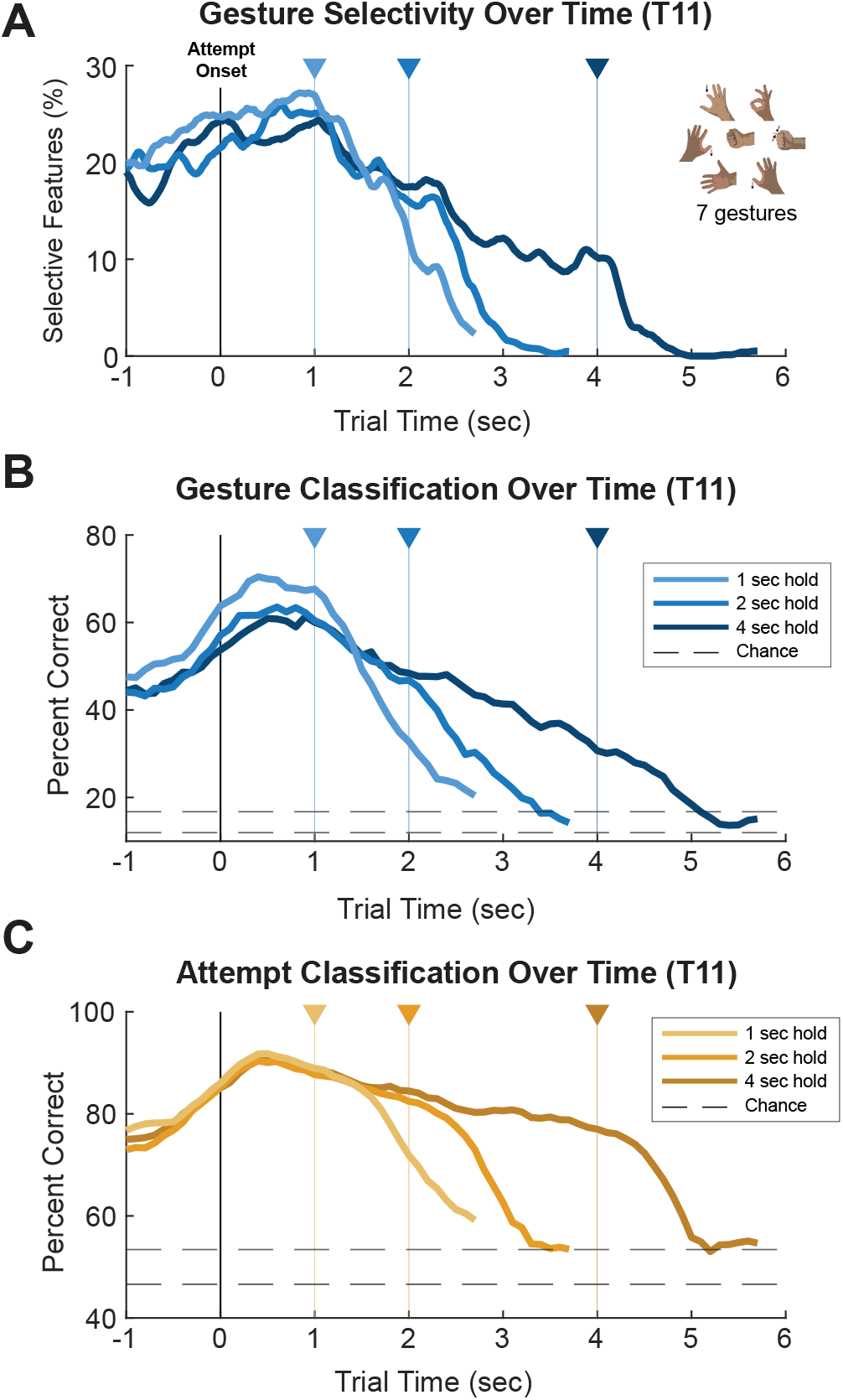
Gesture information in recordings from T11’s motor cortex during the Gesture Hero task. (A) The percent of neural spiking features (TX and SP) selective for gesture type assessed on 300ms windows incremented in 20ms increments over each set of trial durations (1s, 2s, and 4s trials). Features were considered gesture selective in a given time window if they displayed significantly different values across the 7 gesture conditions (KW test, *p <* 0.001). (B) The performance of LDA classification in discriminating between the seven gesture conditions across each trial duration. Cross-validated (5-fold) accuracies were computed on classifiers built on data from 500ms windows stepped in 100ms increments. Dashed lines reflect the 95% chance interval. (C) The performance of LDA classification in discriminating between all gestures and the intertrial period.

When applying the same analysis to the neural signals of T5 performing the Gesture Hero task (with only 3 gestures), we likewise found a significant drop off in gesture selectivity, with about 12% gesture selective features at the end of 1s hold trials, 6% at the end of 2s hold trials, and 3% at the end of 4s hold trials (Fig. S1A). Although gesture classification (LDA) performance assessed over time (Fig. S1B) also showed a decrease throughout T5’s sustained gesture attempts, this relative drop off (a 12.3% decrease from 1s to 4s) was far shallower than T11’s, even when compared with LDA classification of T11’s data evaluated on the subset of gesture trials performed by T5 (index finger down, power grasp, OK, 35.2% decrease from 1s to 4s; Fig. S2B). Curiously, attempt classification of T5 data did not demonstrate a meaningful decrease in accuracy over the trial duration (Fig. S1C), even revealing the presence of a second “peak” shortly *after* the end of sustained gesture trials.

We also assessed the percent of features that were attempt selective (i.e. demonstrated significantly different values during gesture attempts compared to baseline; Wilcoxon Rank Sum, *p <* 0.001) during Gesture Hero trials and found notable differences between T11 and T5 in the neural representations (Fig. 3A-B). Whereas, for T11, the percent of attempt selective features peaked at the beginning of the trial and smoothly declined over time (Fig. 3A), for T5, attempt selectivity exhibited distinct peaks at both the onset and offset of the trial (Fig. 3B).

**Figure 3.**
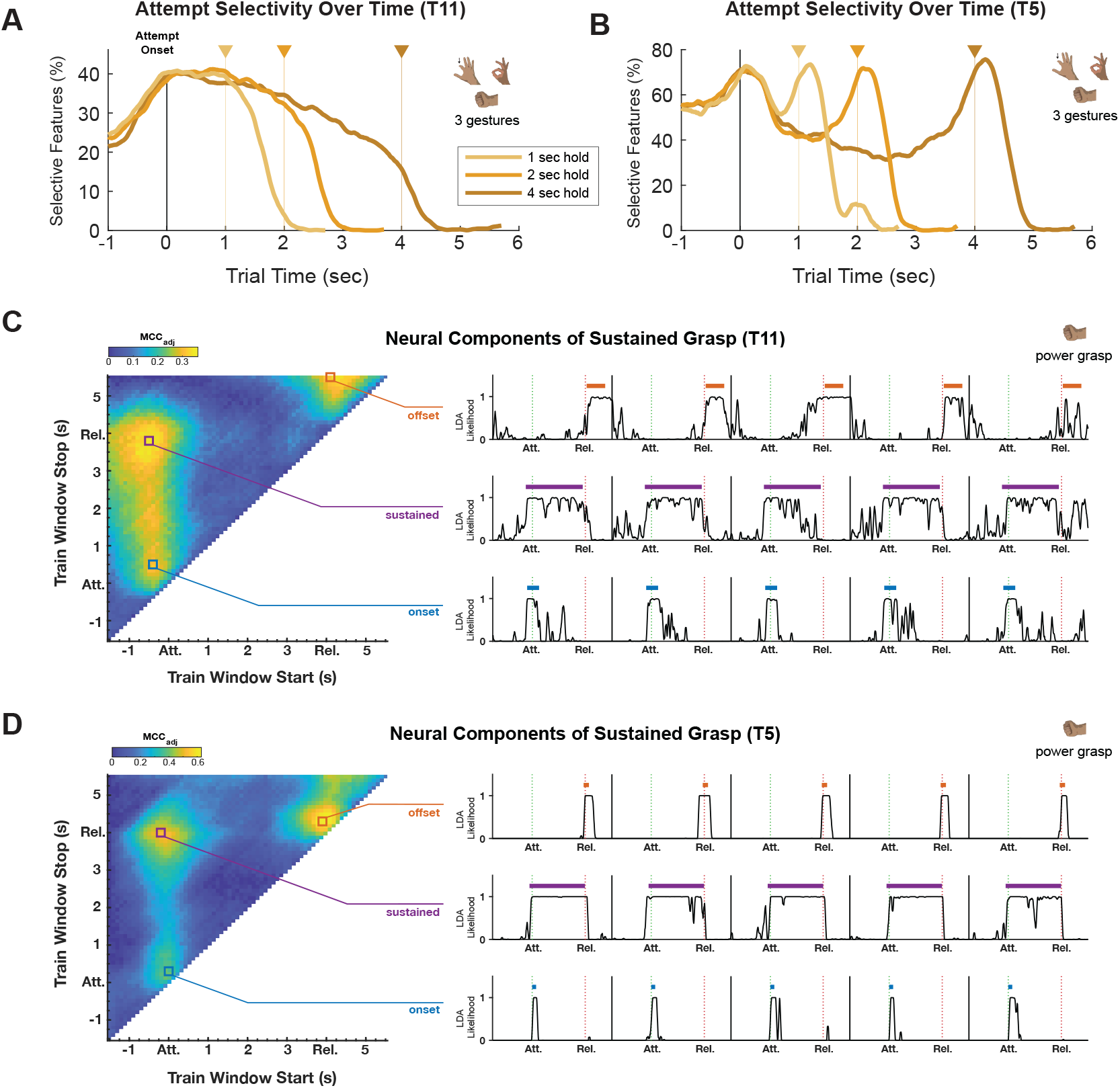
Comparison of Neural Components Between Participants. (A-B) The percent of neural features selective (TX and SP) for *attempt* assessed on 300ms windows incremented in 20ms increments over each set of Gesture Hero trial durations (1s, 2s, and 4s trials). Features were considered attempt selective in a given time window if they displayed significantly different values within that window compared to baseline data (Wilcoxon Rank Sum *p <* 0.001). Baseline data were taken from the 300ms window beginning 1.2s after the end of different duration trials (e.g. attempt selectivity for 1s trials was calculated by comparing the 1s trial data to the baseline windows of the 2s and 4s trials.). Results shown using the subset of 3 gestures (index finger down, power grasp, and OK) for both T11 (A) and T5 (B). (C-D) Results of a grid search using only 4s power grasp trials for T11 and T5 (see Fig. S3 for results from 1s and 2s trials). Each point on the heat plots on the left represent the performance (adjusted Matthews correlation coefficient; *MCC*_*adj*_ ; 5-fold cross-validated) of a binary LDA classifier in differentiating between neural data within and outside of a given time window (“Train Window”). The plots on the right display example LDA likelihood outputs from classifiers built on local maxima from the the grid search process, described by Dekleva *et al* [21] as the “onset transient” (blue), “offset transient” (orange), and “sustained response” (purple). Colored bars above the likelihood traces denote the start and end of the training window corresponding to each local maximum. The Attempt Onset (“Att.”) and Release (“Rel.”; i.e. when the participant was supposed to start and stop attempting a power grip) are denoted by dotted green and red lines, respectively. Note that although there is a distinct “offset” signal, centered at the Release mark, that emerges from the grid search on T5’s data (D), the corresponding local maximum identified in T11’s data is broadly centered over the intertrial period and more likely represents activity specific to the “relax” state assumed by the participant between trials, rather than activity related to the release, or “offset”, of the power grip gesture.

The presence of “onset” and “offset” responses in T5’s neural data resembles previous descriptions of distinct neural activity patterns, or *components*, associated with the onset, offset, and “sustained” periods of single-gesture drag-and-drop trials performed by other iBCI participants [21]. These neural components were identified by performing an exhaustive grid search of all possible trial-aligned time windows, assessing the ability of a binary LDA classifier to differentiate between neural data collected within and outside of each time window [21]. When applying this approach to the 4s Gesture Hero trials, we found peaks in classifier performance (measured by an “adjusted” Matthews correlation coefficient (*MCC*_*adj*_; see [21])) corresponding to onset and sustained components in both T11 and T5, but only data from T5 exhibited a distinct “offset” component (Fig. 3C-D, Figs. S3-S5).

The lack of reliable gesture offset components in T11 suggested that the approach of using two independent classifiers to control the onset and offset of sustained gesture attempts would not generalize well for all participants. We therefore designed a new approach that combines the transient, highly gesture-selective signal present at the beginning of a sustained gesture attempt (Fig. 2A-B) with the more robust, attempt-selective signal (Fig. 2C) present throughout the duration of a held gesture.

### 3.3 The Latch Decoder

Given that the peak in gesture-related information encoded in precentral neuronal activity was noted to occur at the onset of the attempted gesture, we developed a decoding strategy that would “latch” or maintain the initial decoded gesture: the Latch decoder (Fig. 4A). The Latch decoder consists of two component. One component predicts the gesture type (Gesture), and the other predicts if any gesture is being attempted (Attempt). Initially, the Latch decoder’s output (Latch State) directly reflects the Gesture decoder’s state (Gesture State). However, when the Attempt State is true and the Gesture State has been the same for 400ms (*t*_*Latch*_), the Latch State becomes *latched* to the present decoded gesture type until the Attempt State becomes false.

**Figure 4.**
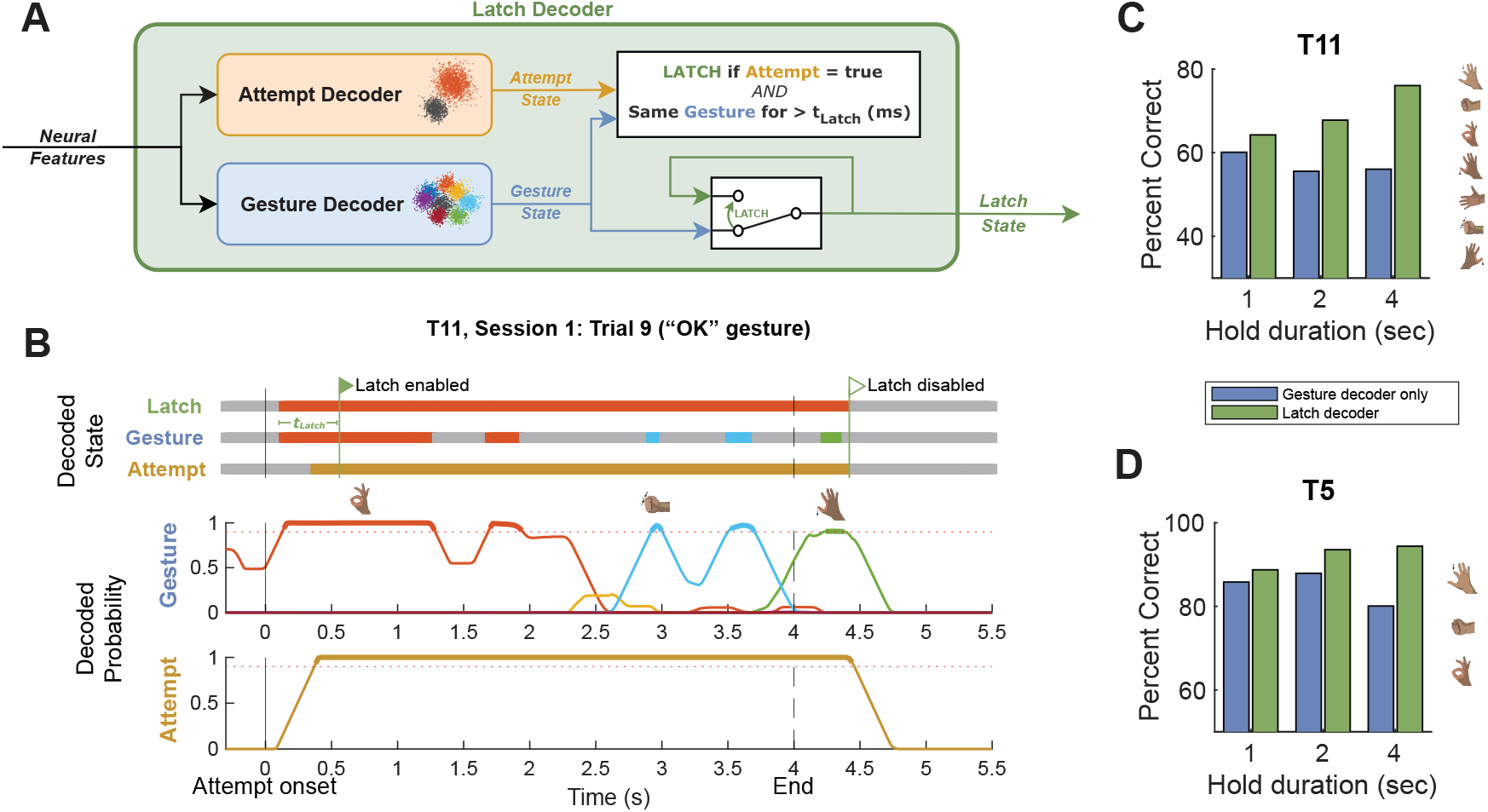
(A) Diagram detailing the Latch decoder pipeline. Selected neural features are input to both the binary Attempt decoder and the multi-class Gesture decoder, which run in parallel. If the Attempt decoder decodes an attempt and the Gesture decoder has decoded the same gesture type for more than 400 ms, then we “latch” to the current decoded gesture type until the attempt decoder no longer decodes an attempt. (B) Decoded states (top) and normalized emission probabilities (bottom) from an example trial from the Gesture Hero task wherein T11 was prompted to hold an “OK” gesture for 4 seconds. Around 200ms after attempt onset, the Latch State and Gesture State both represent the correct gesture prediction. At ∼600ms, this initial gesture prediction is “latched” because the Gesture State has been the same for 400ms (*t*_*Latch*_) and the Attempt State is true. The Latch State returns to the relax/no-action state at ∼4.5s when the Attempt State becomes false. The Latch decoder compensates for multiple potential sources of error, including errors where the gesture is no longer decoded and when other gesture probabilities go above threshold during a hold period. (C-D) Offline cross-validated (10-fold) performance of the Gesture decoder (alone) and the Latch decoder on 1s, 2s, and 4s Gesture Hero trials collected from T11 and T5. Bars represent the mean percent of correctly decoded time steps (20ms steps) during the hold duration (i.e. for what percent of each hold period was the correct gesture decoded). To account for reaction time, only time steps after the first correctly decoded time step were evaluated.

Fig. 4B shows an example of how the Latch decoder operates. The Latch decoder’s state becomes latched ∼600ms after the attempt onset and does not change despite the changes in the Gesture decoder’s output. Its output becomes unlatched ∼4.5 sec into the trial when the Attempt State transitions to false. Using this approach on data collected from the Gesture Hero task, we found that the Latch decoder enabled substantial increases in the percent of correctly decoded time steps compared to using the Gesture decoder alone for both participants (Fig. 4C-D).

Both the Gesture and Attempt decoders used linear discriminant analysis (LDA) in conjunction with a hidden Markov model (HMM), LDA-HMM, as specified in Hosman *et al* [5]. In brief, incoming z-scored features were smoothed with a 100ms boxcar filter and projected to a low dimensional space before class posterior probabilities were computed using LDA. Emission probabilities were then produced via an HMM, smoothed with a 100ms boxcar filter, and thresholded (see Fig. 4B) to determine the “decoded state” output from each decoder.

Whereas the Gesture decoder was trained to differentiate between all gesture classes (including the relax state), the Attempt decoder was trained on the same data, but all gesture classes were relabeled as a single “attempt” class. A regularization parameter (see [25]) empirically set to 0.3 was used when computing the LDA coefficients, and the class means and covariances used the empirical mean and covariances from the calibration data. For the Gesture decoder, the HMM transition matrix was set to match the gesture state transitions of the calibration task. The HMM transition matrix of the Attempt decoder was manually set to be extra “sticky” [26], with on-diagonal values of 1 − 10^−10^, to prevent transient misclassification of the attempt signal. Of the 1,344 recorded features (one TX value, one SP value, and the five lower-frequency LFP bands per channel, 192 channels), the top 400 were selected for classification analysis by identifying all gesture selective (or attempt-selective) features (Kruskal-Wallis, *p <* 0.001) and ranking them according to minimum redundancy maximum relevance algorithm [24]. This method allows for optimal feature selection for each decoder individually. For example, when testing the Latch decoder on 4s hold trials from the Gesture Hero dataset (4C-D), the Gesture decoder utilized TXs, SPs, and higher frequency LFP features, whereas the Attempt decoder used very few TX features and relied more heavily on LFP features (Fig. 5).

**Figure 5.**
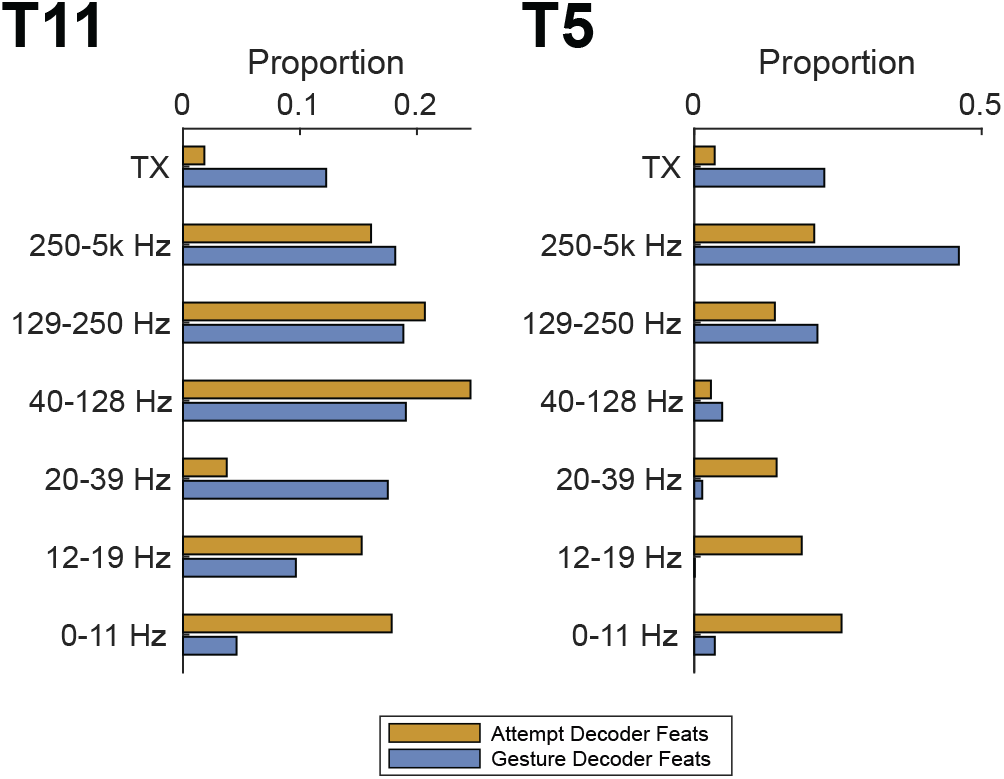
Independent feature selection for Attempt and Gesture decoders. When building LDA-HMM decoders for attempt classification and gesture-type classification, an independent subset of 400 features are selected for each decoder by identifying all gesture (or attempt) selective features and choosing the top 400 features after ranking with a minimum redundancy maximum relevance algorithm [24]. Bars represent the proportion of each type of feature used in the Attempt and Gesture decoders that were built when calibrating a Latch decoder on data 4s sustained gesture trials from the Gesture Hero task (4C-D).

## 4. Multi-Gesture Drag-and-Drop Study

Rarely are sustained gestures and clicks performed in isolation when using a personal computer or tablet. Thus, we designed a multi-gesture drag-and-drop task to assess how well the Latch decoder enables sustained grasp decoding *while* an iBCI participant is also controlling cursor kinematics.

### 4.1 Drag-and-Drop Task

The Drag-and-Drop Task consisted of a 2D center out and return task with four outer targets positioned cardinally from the center target. There were three trial variations present in each data collection block (Fig. 6A, Video S2):

- **Move Only**: Move Only trials represented a simple kinematic task that required the participant to move a circular cursor from the center of the screen to an outer target (Center Out stage), wait for 1-2.5s on the outer target (Wait), and move the cursor back to the center (Return). In order to acquire the outer target, the cursor needed to *dwell* within the target radius for 0.5 seconds.
- **Click**: During Click trials, the participant performed an identical kinematic task as for Move Only trials but was instructed to perform a transient gesture attempt, or “click”, in order to acquire the outer target.
- **Drag**: Drag trials were similar to Click trials, however, once the cursor reached the outer target, the participant was instructed to attempt and hold the cued gesture for 1-2.5s and then continue holding the gesture while moving (i.e. dragging) the cued gesture icon from the outer target location back to the center. Drag trials contained an additional 1s “Hold” stage at the end wherein the participant continued attempting the gesture without kinematic movement.

**Figure 6.**
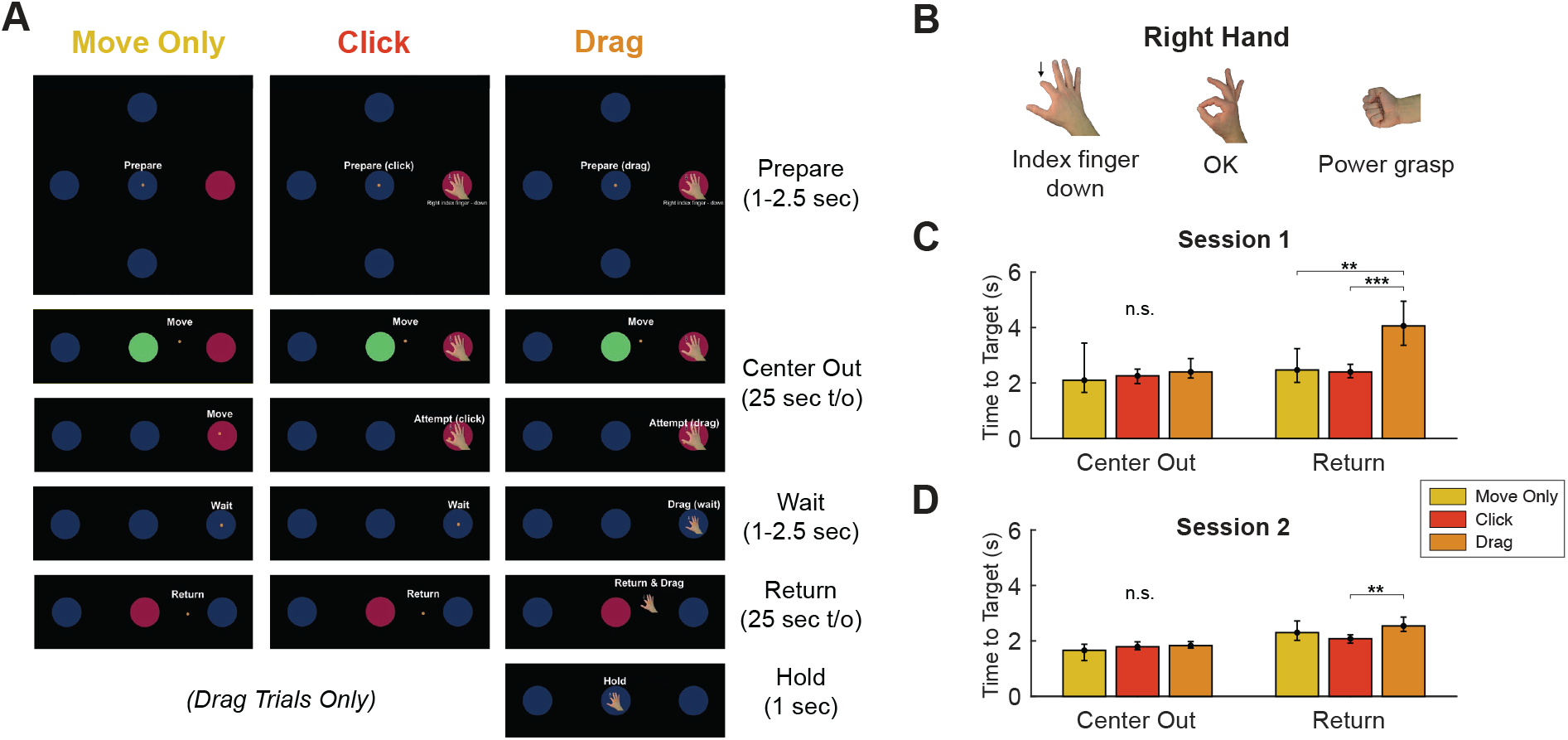
The Multi-Gesture Drag-and-Drop Task. (A) Screenshots (cropped) from example trials where the participant acquired rightward targets under closed-loop two-dimensional iBCI control of a (small, orange) cursor. The task consisted of three trial variations: Move Only (no gesture decoding required), Click (transient gesture attempt needed to acquire outer target), and Drag (sustained gesture attempt required to acquire target and “drag” the gesture icon to the center target). Each trial consisted of a combination of instructed delay stages (Prepare, Wait), self-timed stages with timeout (t/o) limits (Center Out, Return), and fixed duration stages (Hold; for Drag trials only). (B) Three gestures were used during Click trials and Drag trials of the the Drag-and-Drop Task. (C-D) Time to target results during the Center Out and Return stages for each trial variation in T11’s first session (C) and second session (D) performing the task. All attempts were limited to a 25s timeout period. Bar heights represent the median time to target value for each trial variation. Error bars represent 95% confidence intervals. Horizontal bars with asterisks denote significant differences between trial types (Wilcoxon Rank Sum; *, *p <* 0.05; **, *p <* 0.01; ***, *p <* 0.001; n.s., *p >* 0.05).

Each trial began with a random 1-2.5s “Prepare” stage (Fig. 6A) wherein an outer target was cued by changing from blue to red. In Click trials and Drag trials the outer target was also overlaid with an image of one of the three gestures that they were supposed to perform once reaching the target (Fig. 6B). The words “Prepare (drag)”, “Prepare (click)”, or “Prepare” were visible above the cursor during this stage. Thus, the participant had information on the movement direction, the gesture (if applicable), and the trial type he was about to perform from the beginning of the trial. To provide additional guidance during the task, the words “Move”, “Attempt”, “Drag”, “Return”, and “Hold” were presented above the cursor (see Fig. 6A) to instruct the participant during each stage of the trial. During Drag trials, the gesture icon would shrink in size and follow the cursor to represent “dragging” of the icon when the correct gesture was being decoded. If the decoder did not decode the correct gesture, the gesture icon would be “dropped” (i.e. the icon would return to original size and cease moving along with the cursor). Both the Center Out and Return stages had timeouts of 25 seconds.

Participant T11 performed the Drag-and-Drop Task during two sessions, each with a total of 11 blocks. Each block contained 4 Move Only trials (one for each direction), 12 Click trials (one for each direction/gesture combination), and 12 Drag trials (for each direction/gesture combination). The first four blocks were used to calibrate the decoders (steady state Kalman filter [27]) for 2D kinematic decoding and the Latch decoder for gesture decoding) and the seven subsequent blocks were treated as assessment blocks.

All trials in the first block were “open loop” (OL) trials, meaning the computer displayed idealized performance of the Drag-and-Drop task while the participant imagined movements corresponding with the task. For this task, T11 imagined translating his right arm in a horizontal plane to create 2D movement of the cursor and performing the cued gestures with his right hand. For blocks 2-4, kinematic control was gradually given to the participant through step-wise reduction of error attenuation (EA). EA reduces the decoded cursor velocity in the error direction by a factor corresponding to how much assistance we want to provide to an incompletely calibrated kinematic decoder. For example, EA of 0.5 would reduce the decoded velocity component perpendicular to the cursor-target vector by half. Block 2, 3, and 4 had EA values of 0.5, 0.3, and 0.0, respectively, with new Kalman filters generated after each block that incorporated previous blocks’ data (see Video S2). For all calibration blocks, gesture decoding was inactivated (i.e. remained completely OL). All seven assessment blocks were performed in “closed loop” (CL), under full participant control.

### 4.2 Kinematic Performance

Within each trial of the Drag-and-Drop task there were two kinematic tasks: moving the cursor from the center to an outer target (Center Out stage) and returning from the outer target (Return). Despite contextual differences between Move Only, Click, and Drag trials, the instructed task during the Center Out stage was functionally the same between trial types. Thus, as expected, target acquisition times during this stage were not significantly different across trial types (Kruskal-Wallis (KW), *p* = 0.265, Fig. 6C-D). However, trial timeouts were more common during Drag trials (2.4%) than Click trials (1.1%) and Move Only trials (0.0%).

During the Return stage, whereas Move Only and Click trials were functionally the same, Drag trials required the participant to perform the same kinematic task while simultaneously holding a gesture. T11’s median target acquisition times were significantly greater during the Drag trials (3.16s) than for Click (2.22s; Wilcoxon Rank Sum (RS), *p* = 3.92e-07), and Move Only (2.30s; RS, *p* = 0.002) trials. However, this difference was far more noticeable during the first session (Fig. 6C) than the second session (Fig. 6D), suggesting a learning effect. Moreover, when considering Session 2 trials alone, Drag (Return) trial durations were not significantly different from Move Only trials (RS test, *p* = 0.186).

### 4.3 Latch Decoder Performance

During Drag trials of the Drag-and-Drop task, T11 used the Latch decoder to select and maintain selection of gesture icons across three trial stages: Wait, Return, and Hold. We evaluated the occurrence of gesture decoding errors during these epochs and compared performance to what would have occurred if the output of the Gesture decoder component was used on its own. Here, an error is considered an incorrect gesture decode, including a no gesture decode. Note that for the Wait period, to account for variance in reaction times, we only consider steps after the decode onset (correct or incorrect). If there was no decode during the Wait period, we counted the entire period as incorrect.

Using the Latch decoder, T11 completed 73% of Wait epochs, 79% of Return epochs and 86% of Hold epochs without a gesture decoding error. By contrast, if the output of the Gesture decoder was used on its own, only 41% of Wait epochs, 3% of Return epochs, and 15% of Hold Epochs would have been completed without error (Fig. 7A). Specifically, during Wait, Return, and Hold epochs, the Latch decoder, on average, output the correct gesture decode for 92%, 96%, and 93% of the duration of each epoch, respectively. Meanwhile, the Gesture decoder output reflected the correct gesture 86%, 61%, and 56% of the duration of Wait, Return, and Hold epochs, respectively (Fig. 7B).

**Figure 7.**
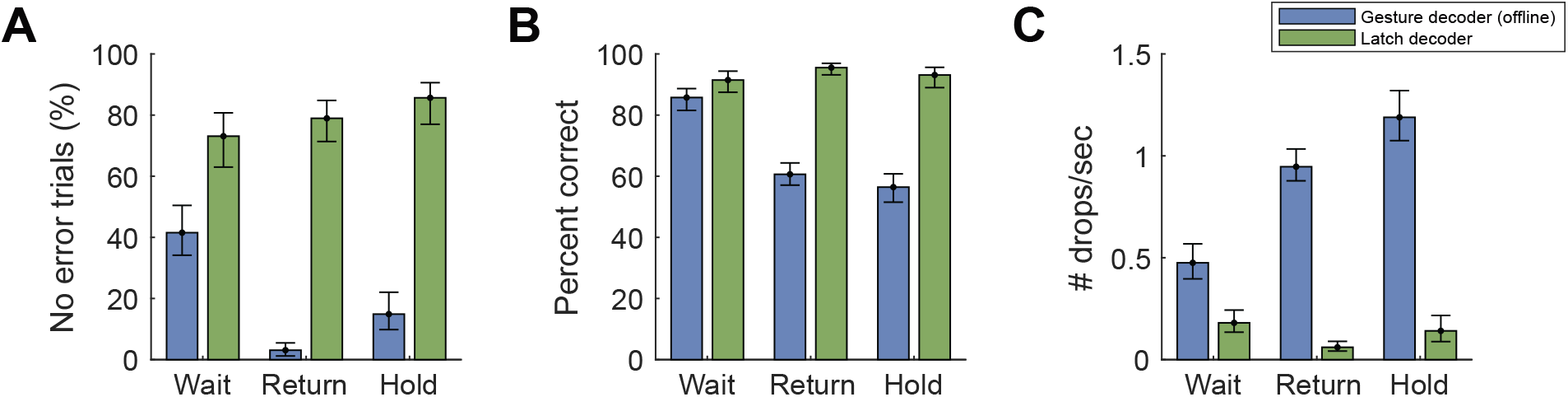
Performance of the Latch decoder compared to the Gesture decoder on its own (simulated offline) during Drag trials of the Drag-and-Drop Task performed by T11. (A) The percent of trial epochs without any error. Estimated each block and showing means per trial epoch (Wait, Return, Hold) with 95% CI error bars. (B) Percent correctly decoded gesture across each individual trial. Mean across all trials with 95% CI error bars. (C) The number of dropped events per second. Mean across all trials with 95% CI error bars.

The Latch decoder showed particular promise in preventing unintended drop events during the task, especially during the Return and Hold periods when the gesture attempt was being sustained beyond 1 or 2 seconds. Using the Latch decoder, T11 averaged 0.2, 0.1, and 0.1 drop events per second during the Wait, Return, and Hold periods, respectively. Using the Gesture decoder alone would have yielded 0.5, 0.9, and 1.2 drops per second, respectively.

## 5. Discussion & Conclusion

This study demonstrates that the Latch decoder can provide iBCI participants with significant improvements in the ability to “drag and drop” icons using multiple gestures on a computer screen. The Latch decoder improves upon existing multi-gesture LDA-HMM classification of sustained gestures by taking advantage of the relatively higher neural discriminability of the overall “attempt” signal to “latch” to decoded gesture types inferred at the beginning of a movement period, capturing the phasic component of the neural response. Other approaches that were considered for accommodating decreasing signal quality during the hold period involved increasing the smoothing of neural features, increasing the time window on which the decoder was evaluated, or making the HMM transition matrix of the Gesture decoder more “sticky” [26]. However, a drawback of these methods was that they introduced significant delays in the decoding of gesture onsets that would cause the experience of using the decoder to be unacceptably sluggish. A key benefit of the Latch decoder is that the accuracy and latency of decoding transient gesture attempts (or clicks) is unaffected because decoding of sustained gesture attempts is provided by a separate, parallel process that is only activated 400ms after gesture onset.

Existing toggle-based approaches that use transient gesture attempts to drive changes in the controlled effector (e.g. hand flexion to close a robotic hand and hand extension to open the hand) require at least two distinct actions to control each degree of freedom. This can be seen as unintuitive for the user as the “return” of many held postures can be more readily interpreted as a “release” of a sustained motor attempt rather than activation of antagonistic muscle groups. Furthermore, this method potentially underutilizes 50% of the available motor repertoire, which could be used to drive additional degrees of freedom. Previous work describing the onset and offset of attempted gestures as distinct neural events in human motor cortex [21] presents a possibility that the initiation, maintenance, and release of an action like a mouse click could be be controlled by a single gesture. However, like the toggle-based approaches, a decoder that classifies the onset and offsets of individual gestures requires two classifier states to control each gesture, and it is unclear how well this method might scale to decoding of multiple sustained gestures states.

The present study revealed a difference in the neural representations of gesture ‘offsets’ between participant T5 and T11, with a clear offset response recorded in T5 but not in T11 (Fig. 3, Figs. S3-S5). This difference could be attributed to slight differences in how each participant approached the task. Despite being given the same instructions, it is possible that T5 employed a more deliberate strategy to end gestures (e.g. envisioning opening the hand), while T11 took a more passive approach (e.g. simply relaxing when a gesture was released). Inter-participant differences in internal imagery are expected sources of variability in iBCI studies that ultimately can be influenced by difficult to control variables such as previous research activities performed using the iBCI or even prior experiences using computers before becoming paralyzed. Furthermore, despite T5 and T11 having arrays implanted in anatomically similar locations in the dorsal precentral gyrus, there is no guarantee that recorded signals represent activity from functionally comparable neural populations. Slight variations in array placement could have yielded a strong offset component in T11 and no offset in T5. It is important to note that the latch decoder was designed to be effective for both participants evaluated in the study.

Preserving grasp-related decoding during translation is a recognized challenge [15]. Results from the multi-gesture drag-and-drop task indicate that the Latch decoder provides improved gesture decoding not only during isolated gesture holds, but also during simultaneous 2D kinematic control of the cursor (Fig. 7A-C). Compared to Click and Move Only trials, Drag trials took longer to complete, but this is also a behavior observed during able-bodied drag-and-drop studies [28] and could be due to cognitive factors unrelated to decoder performance [29]. Notably, T11’s performance of drag trials greatly improved from Session 1 (Fig. 6C) to Session 2 (Fig. 6D), suggesting longer transport times could be overcome with practice. Further work is needed to understand the neural representations of movement interactions during simultaneously controlled degrees of freedom during iBCI control.

The ability to hold specific gestures over longer time periods will expand the ways in which people using an iBCI can interact with screen-based devices like computers and tablets. Perhaps the most direct application of the Latch decoder will be on enabling more intuitive control of anthropomorphic robot arms or one’s own arm using functional electrical stimulation [19, 30]. However, further research will be needed to understand how well our approach translates to these other contexts.

## Data Availability

All data produced in the present study are available upon reasonable request to the authors.

## Acknowledgments

The authors would like to thank participants T11, T5, and their families and care partners. We also thank Beth Travers, Dave Rosler, and Maryam Masood for their contributions to this research.

## Disclaimer

The content is solely the responsibility of the authors and does not necessarily represent the official views of the NIH, the Department of Veterans Affairs or the US Government. Caution: Investigational Device. Limited by Federal Law to Investigational Use.

## Supplementary Material

**Figure S1.**
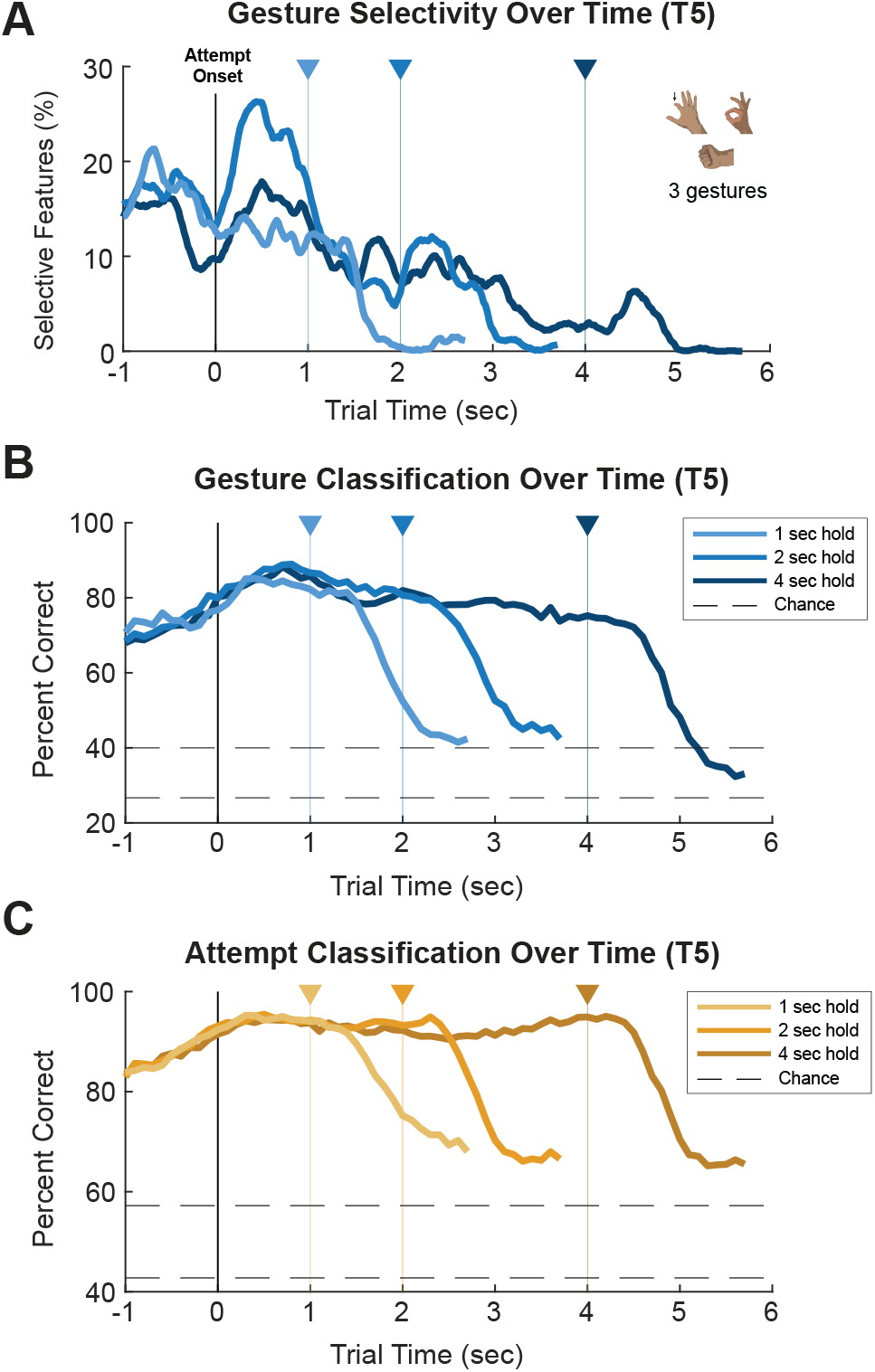
Gesture information in recordings from T5’s motor cortex during the Gesture Hero task. See Fig. 2 caption for details. Note that T5 performed the Gesture Hero task with only 3 gestures: index finger down, power grasp, and OK.

**Figure S2.**
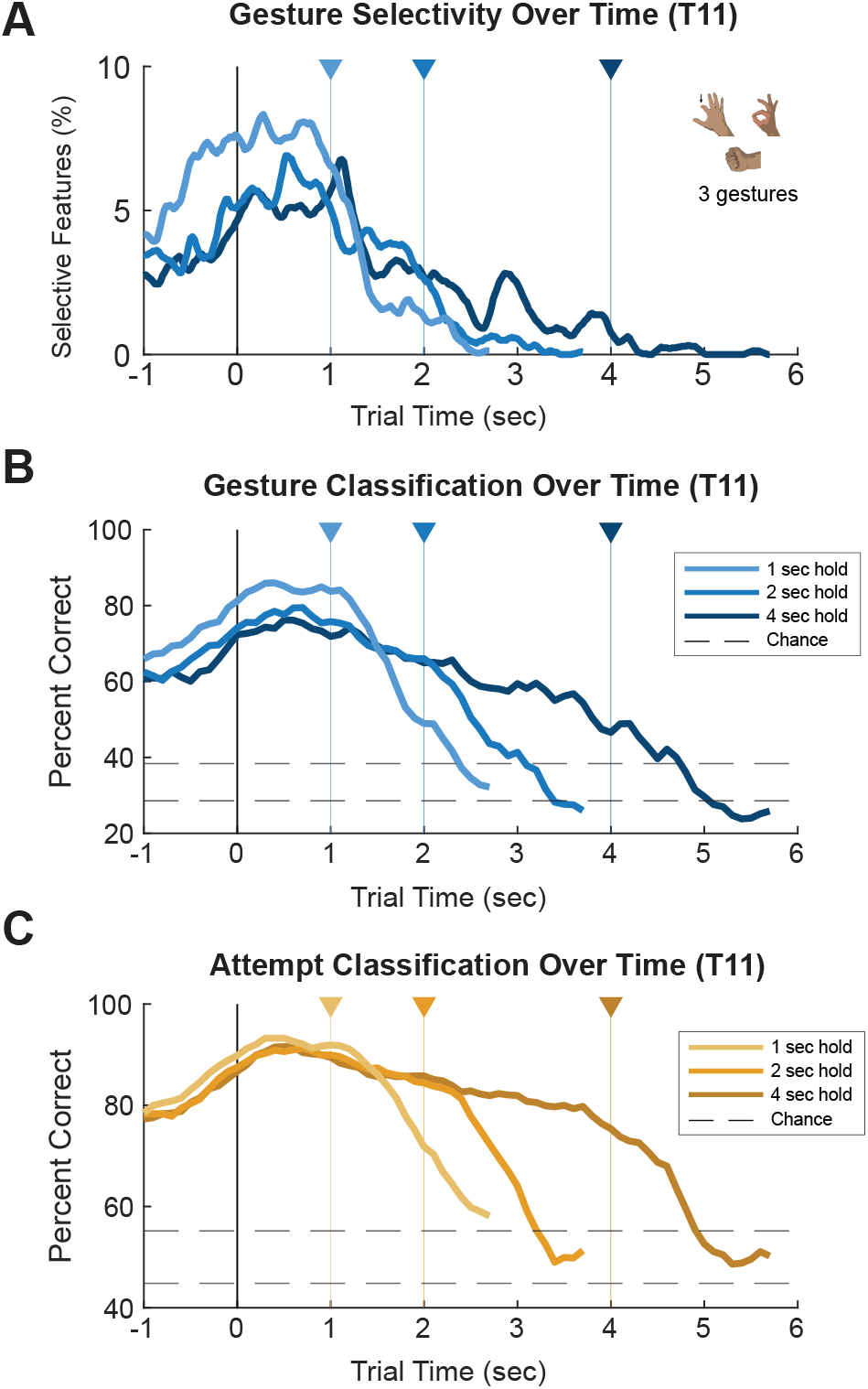
Gesture information in recordings from T11’s motor cortex during the Gesture Hero task using three gestures. Same as Fig. 2 (main text) but evaluated on the subset of three gestures used in T5 sessions to serve as better comparison with T5 results (Fig. S1).

**Figure S3.**
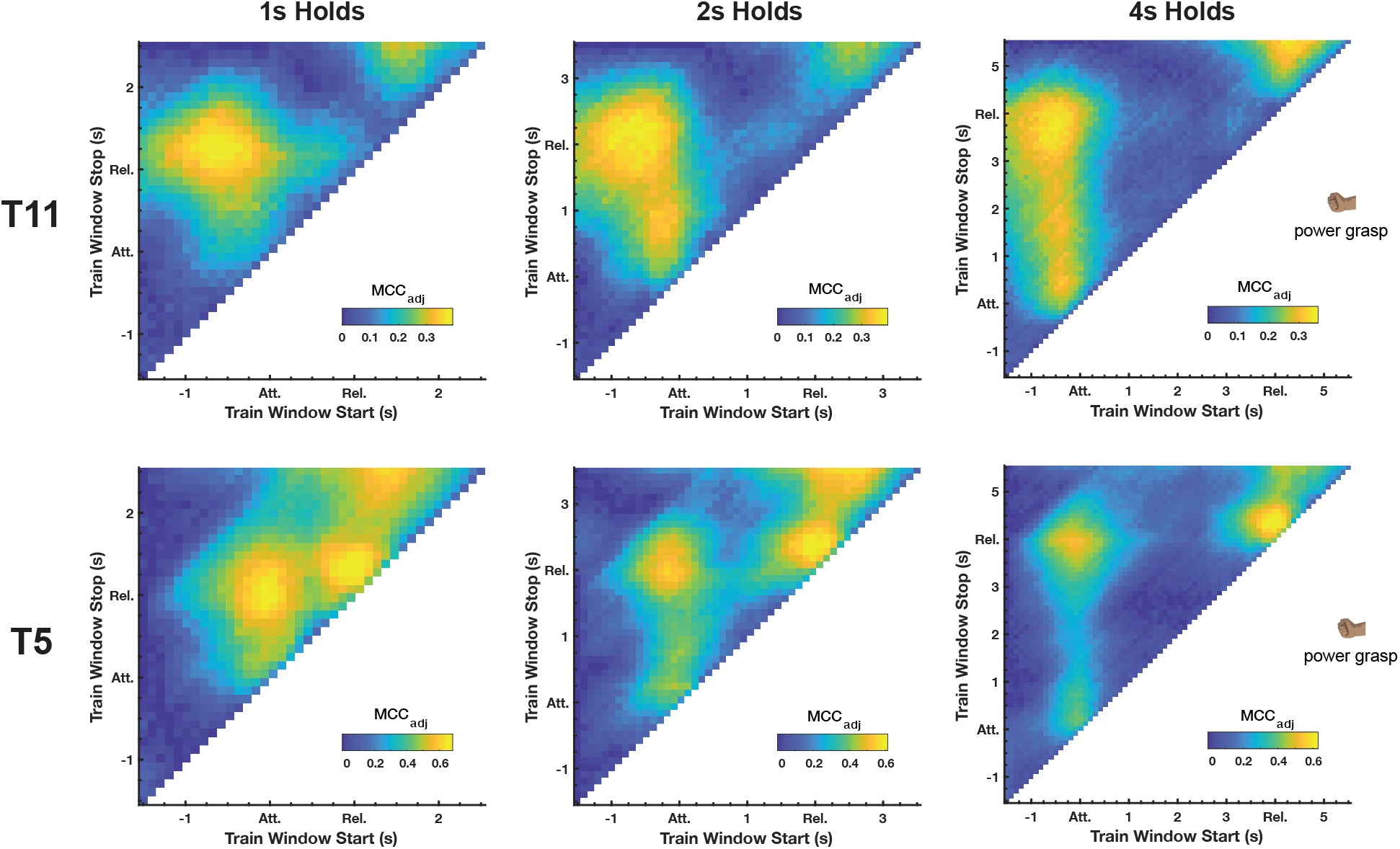
Grid search results for Gesture Hero power grip trials across all hold durations. See Fig. 3 legend for details. Heat plots indicate the presence of a clear “offset” component across all hold durations in T5 data that is not present in T11 data. For both participants, the gesture “onset” component appears to overlap with the “sustained” component in the 1s hold trials.

**Figure S4.**
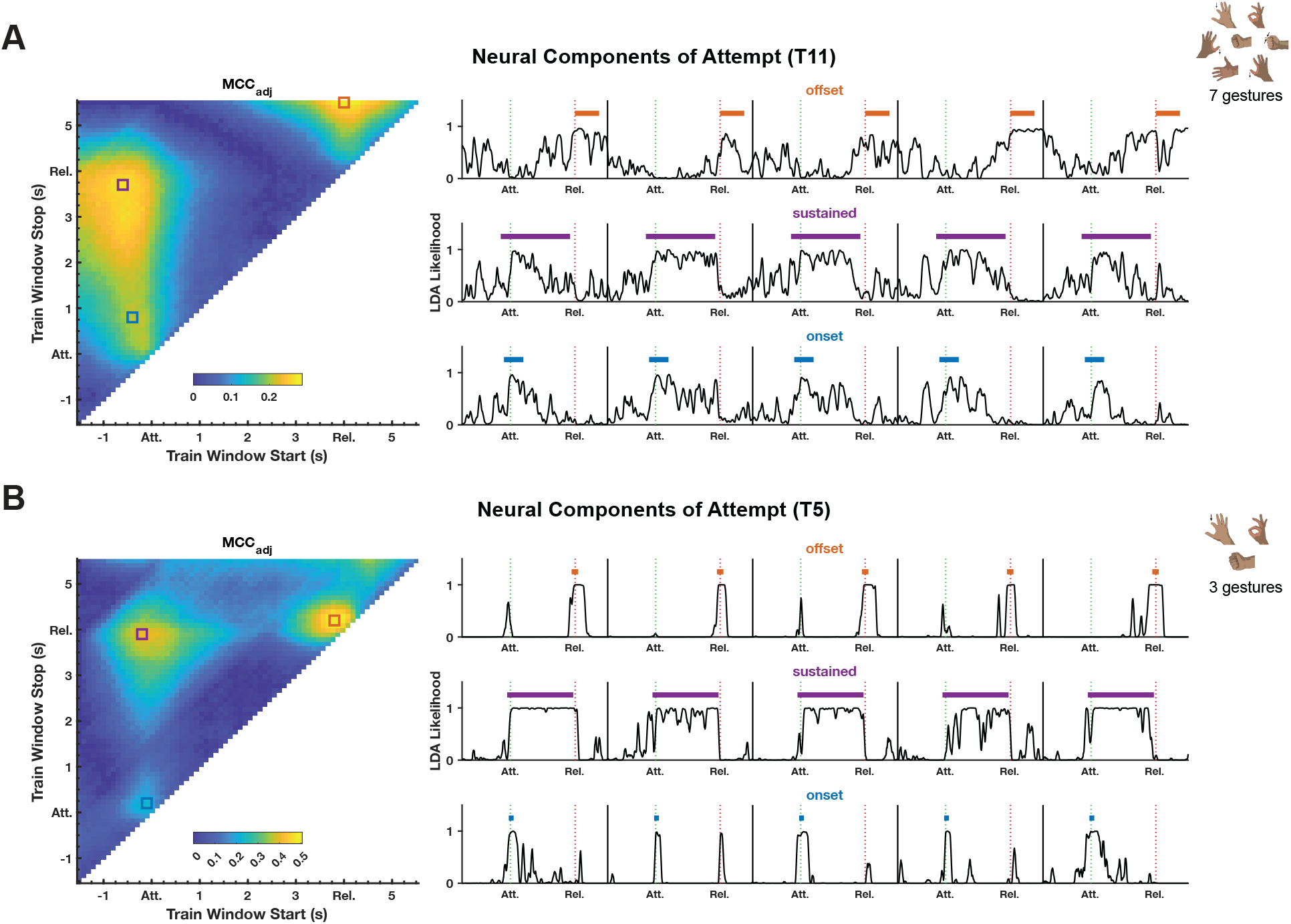
Neural components of “Attempt”. Grid search analyses done on *all* gestures performed during the Gesture Hero task. Similar to Fig. 3C-D, but data from all 4s hold trials are included in generating the binary LDA classifiers for differentiating between neural data within and outside of the training windows swept in the analysis. Like results in Fig. 3C-D, T11’s “offset” component (A) appears centered on the intertrial period, whereas T5’s “offset” component (B) is well-aligned with the gesture release. It is notable that the likelihood traces of both the “onset” and “offset” components in T5 show peaks around both the Attempt and Release periods of the gesture, suggesting that a generalized gesture attempt onset decoder may be prone to false positives around attempt offset (and vice versa).

**Figure S5.**
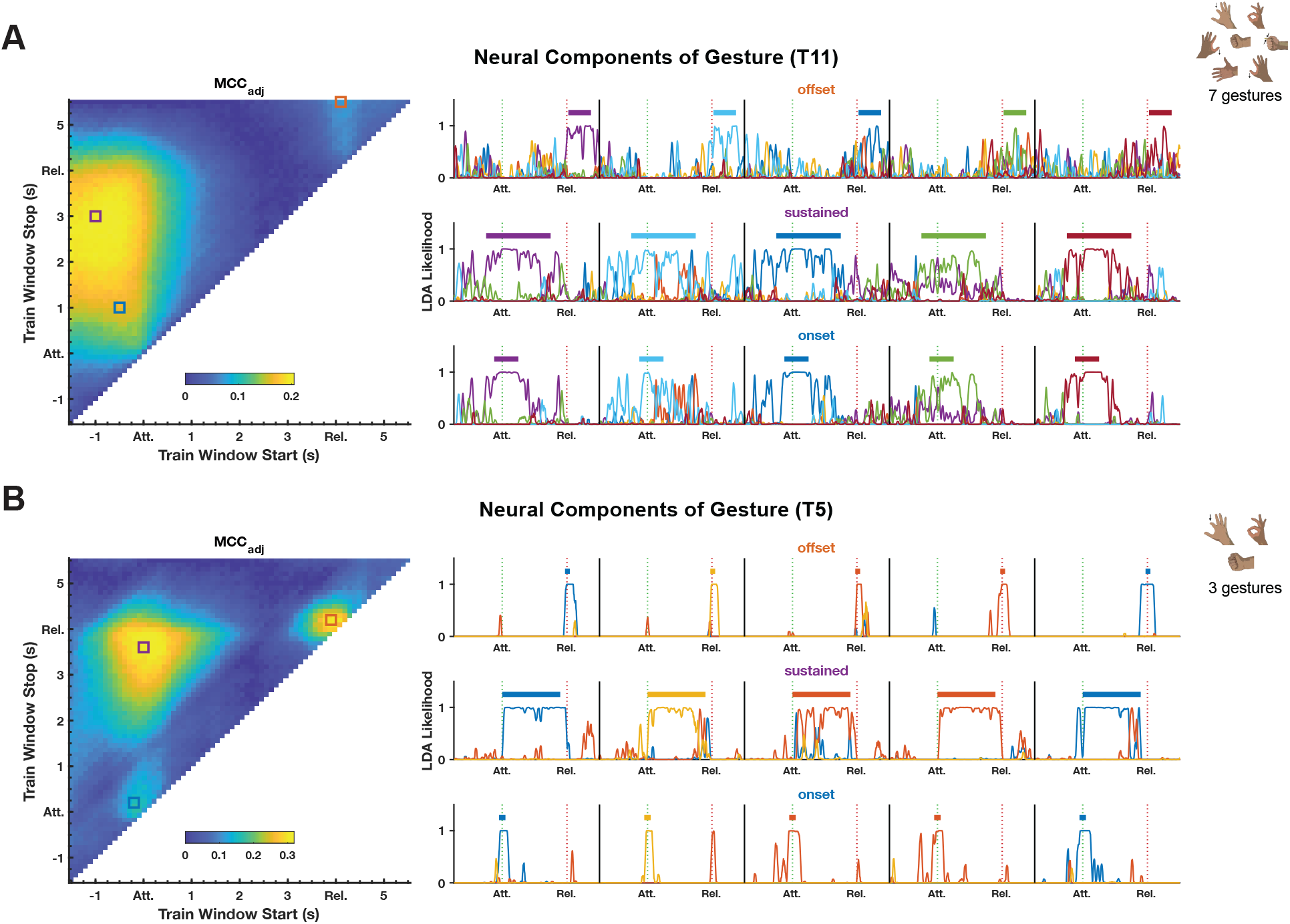
Neural components of “Gesture”. Grid search analysis using a multi-state LDA. Similar to Fig. SS4, but instead of generating binary LDA classifiers that considered all gesture trials as a single class, we evaluated the performance of an LDA classifier to differentiate between all gesture states while sweeping the location of the training window. Results for T11 (A) represent performance of an 8-class LDA (7 gestures + no action) and results for T5 (B) represent performance of a 4-class LDA (3 gestures + no action). Here, the colors of the horizontal bars above the likelihood traces correspond to the gesture that was cued for that trial. Normalized likelihood traces for all gestures (not including the “no action” state) are plotted.

